# Intrapersonal predictors of internalized stigma among school going adolescents living with HIV in southwestern Uganda

**DOI:** 10.1101/2022.06.01.22275873

**Authors:** Joseph Kirabira, Scholastic Ashaba, Allain Favina, Samuel Maling, Denis Nansera, Brian C. Zanoni

**Affiliations:** Department of Psychiatry, Busitema University Faculty of Health Sciences, Mbale, Uganda; Department of Psychiatry, Mbarara University of Science and Technology, Mbarara, Uganda; Emory University School of Medicine, Departments of Medicine and Pediatric Infectious Diseases, Atlanta, Georgia, USA; Children’s Healthcare of Atlanta, Atlanta, USA; Rollins School of Public Health, Department of Global Health, Atlanta, USA

**Author notes:** Corresponding author (JK).

**Keywords:** HIV stigma, self-efficacy, empowerment, resilience, locus of control, Uganda

## Abstract

**Background:** HIV is one of the most stigmatized conditions globally significantly affecting the quality of life of people living with HIV. Stigma particularly affects adolescents living with HIV (ALHIV) due to challenges associated with developmental stage including physical and psychological changes and the need to build peer relationships. The effect of intrapersonal factors including resilience, health locus of control, self-efficacy, and empowerment on HV stigma among ALHIV in Uganda has not been studied. This study aimed at assessing the association between internalized HIV stigma and resilience, health locus of control, coping self-efficacy and empowerment among ALHIV.

**Methods:** We conducted a cross-sectional study between August and October 2020 among 173 adolescents who were attending the HIV clinic at Mbarara Regional Referral Hospital. We measured HIV stigma (the internalized AIDS Related Stigma scale), health locus of control (the 18-item Form C version of the Multi-Dimensional Health Locus of Control measure), resilience (25-item Resilience Scale), self-efficacy (the coping self-efficacy scale), and empowerment (the 28-item empowerment scale). Linear regression models were run to determine the association between HIV stigma and intrapersonal factors and adjusted for sociodemographic characteristics.

**Results:** The median age of participants was 16 (IQR 15-18) years and the median HIV stigma score was 3 (IQR 2-4). There was a negative correlation between HIV stigma and internal health locus of control (b= -0.08, p<0.001), resilience (b= -0.03p<0.001) and coping self-efficacy (b= -0.02, p<0.001) while empowerment score was positively correlated (b= 0.05, p=0.003). However, after mutually adjusting for the intrapersonal factors (resilience, internal locus of control, empowerment and coping self-efficacy), and sociodemographic characteristics, only internal health locus of control (p=0.008) and coping self-efficacy (p<0.001) remained significantly associated with HIV stigma.

**Conclusion:** Internal health locus of control, resilience, coping self-efficacy and empowerment are strong intrapersonal predictors of internalized stigma among ALHIV. Designing interventions focusing on strengthening these factors among the adolescents may be a significant step in the fight against internalized HIV stigma.

## Introduction

By the end of 2020 there were 170,000 young adults aged 15-24 years living with HIV in Uganda constituting 12% of the 1.4 million people living with HIV in the country (Ministry of Health, 2021). Although availability of and access to antiretroviral therapy (ART) has enabled children born with HIV to survive into adolescence and young adulthood HIV remains a leading cause of mortality among adolescents in sub-Saharan Africa (UNAIDS, 2018). HIV among adolescents is often characterized by high rates of loss to follow up, poor adherence to ART and high rates of vilorogic failure contributing to high mortality rates compared to young children and adults (Mekuria et al., 2015; Zanoni et al., 2016). Despite these challenges, HIV care for adolescents has focused more on access to and adherence to ART neglecting the evolving social and psychological needs related to normal development challenges that complicate HIV care in this population (Folayan et al., 2014; Judd et al., 2016).

Many ALHIV report experiencing stigma and rejection among their peers and the community (Ashaba et al., 2018; Ashaba et al., 2019; Cervia, 2013) and HIV stigma remains a major barrier to HIV care in this age group affecting ART adherence and retention in care (Martinez et al., 2012; Vreeman et al., 2009). HIV stigma and discrimination also impact access to and enrollment into HIV care among ALHIV due to the fear of rejection and loss of respect among peers (Ashaba et al., 2019; Nabukeera-Barungi et al., 2015). Attending boarding schools (Birungi et al., 2011), where living arrangements may not provide privacy in relation with HIV medications, increases the risk of unintended exposure of their HIV status and the experiences of HIV stigma (Inzaule et al., 2016; Mutwa et al., 2013). ALHIV in boarding schools may resort to hiding their HIV medications while others may fail to honor their HIV clinic appointments to avoid being identified as living with HIV interfering with their ability to adhere to treatment (Ashaba et al., 2019; McHenry et al., 2017). For others, rigid school routines do not offer the needed flexibility to enable adolescents to take their medicines at designated times (MacCarthy et al., 2018; Zanoni et al., 2019).

Intrapersonal factors like self-efficacy, health licus of control, empowerment and resilience are known to influence the perception of HIV (Seghatol-Eslami et al., 2017). Self-efficacy has been associated with positive health related outcomes (Steele et al., 2013) and documented as a major factor influencing stigmatization among people with chronic medical conditions (Johnson et al., 2007; McCann et al., 2008). Additionally, resilience which is the ability of an individual to cope in the presence of adverse situations (Zimmerman, 2013) has been associated with less feelings of stigma (Gottert et al., 2019). Internal health locus of control (ILOC) (the belief that health is in one’s control) has been found to influence quality of life and reduce feelings of stigmatization among people living with HIV (Hua and Vosvick, 2008; Mostafavian et al., 2018). On the other hand, empowerment which is associated with development of self-efficacy and health locus of control has been noted to improve resilience (Brodsky & Cattaneo, 2013; Shogren et al., 2014). Improving self-efficacy and internal health locus of control are likely to buffer the effects of internalized HIV stigma and consequently improve ART adherence (Seghatol-Eslami et al., 2017; Turan et al., 2017).

Although multiple studies indicate that coping self-efficacy, resilience, empowerment and health locus of control influence HIV stigma, information on the interaction between these predictors among ALHIV is sparse (Denison et al., 2015; McHenry et al., 2017). This is especially crucial for school going adolescents whose living conditions put them at an increased risk of internalized HIV stigma and poor adherence to medicines (Mutwa et al., 2013). Therefore, this study aimed to understand the intrapersonal predictors ofinternalized HIV stigma among school going ALHIV southwestern Uganda.

## Methods

### Study setting

We conducted the study at the HIV clinic attached to Mbarara Regional Referral Hospital (MRRH) in Mbarara city. Mbarara city is 270 km from Uganda’s capital, Kampala, and it is the area’s commercial hub, with a population of 195,013 (Uganda Bureau of Statistics, 2014). Most of the people accessing care in the HIV clinic at MRRH live in rural areas outside the city majority of whom rely on substance agriculture, animal husgandry and local trading to earn a living with documented challenges of water and food insecurity (Mushavi et al., 2020; Tsai et al., 2016). By 2020, HIV prevalence in Mbarara was estimated at 13% among those aged 15-49 years which is higher than the national prevalence of 5.8% (UNAIDS, 2021). The HIV clinic at MRRH offers HIV care services to both adults and children free of charge. To date, the pediatric and adolescents care section has cared for over 3,000 children and adolescents living with HIV.

### Study participants

between August and October 2020, we recruited 173 adolescents who were aged 13-19 years, fully aware of their HIV status (HIV status fully disclosed to them), attending school, living within 60 km of the HIV clinic, and were willing to provide assent and/or consent to participate in the study. We used purposive sampling to ensure a diverse range of participants (e.g., boarding school vs. day school). We excluded ALHIV who were not fully aware of their HIV status, despite getting antiretroviral medications; those who were not physically strong enough to remain present for the duration of the questionnaire administration; and those who had difficulty fully understanding the interview questions due to cognitive impairments, as assessed clinically in consultation with a licensed Ugandan psychiatrist.

### Sampling procedure

We used consecutive sampling technique to recruit participants for the interview. Participants were assessed by a clinician in the HIV clinic for eligibility before referring them to the research assistant (RA) for enrollment. Those who met the inclusion criteria were given details about the study by the RA before they provided assent and/or consent to participate in the study. Enrollment was done within the HIV clinics after eligible participants completed all the procedures concerned with HIV care. Interviews were conducted in a private room within the HIV clinic to ensure privacy and confidentiality.

### Data collection

We used an aniterviewer adminsitred questionnaire to collect information on sociodemographic characteristics including age, sex, level of education, type of school (day versus boarding), and who they lived with. We assessed for food insecurity using a single question (did you have enough food at home in the past month?). We also collected information on internalized HIV stigma, empowerment, resilience, coping self-efficacy, and health locus of control using tools described below. The main outcome variable was internalized HIV stigma while the main predictor variables were resilience, empowerment, health locus of control and coping self-efficacy.

### Study measures

#### Internalized AIDS-Related Stigma Scale

The Internalized AIDS-Related Stigma Scale (IARSS) is a six-item scale that assesses internalized stigma. It was created for usage in a group of individuals living with HIV from the United States, South Africa, and Swaziland (Kalichman et al., 2009). It is one of the most extensively used stigma measurement scales in the field and has been validated for use in the Ugandan context (Pantelic et al., 2015; Tsai et al., 2013).The IARSS items are focused on self-blame and concealing one’s HIV status. Examples include “it’s difficult to tell people about my HIV infection” and “I’m ashamed to be HIV positive”. Each question is scored on a binary response scale (agree/disagree), and the scale’s final score is computed as the sum of the items’ scores, with higher scores indicating greater internalized stigma. The scale has been used among ALHIV in the current study setting with a Cronbach’s alpha of 0.75 (Ashaba et al., 2018)

#### Coping Self-Efficacy scale (CSES)

Coping self-efficacy was measured using the Coping Self-Efficacy scale (CSES;) (Chesney et al., 2006). The CSES measures participants’ perceived self-efficacy in coping with psychological challenges and threats. Respondents are asked, “When things aren’t going well for you, or when you are having problems, how confident or certain are you that you can do the following?” The questionnaire then lists 13 coping behaviors that tap three distinct dimensions of adaptive coping: problem-focused coping (e.g., “Think about one part of the problem at a time”), emotion-focused coping (e.g., “Take your mind off unpleasant thoughts”), and social support (e.g., “Get emotional support from friends and family”). Respondents endorse their confidence in carrying out these behaviors on an 11-point Likert scale, ranging from 0 (“Cannot do at all”) to 5 (“moderately certain can do”) to 10 (“certain can do”). This tool has been used among adults living with HIV (Johnson et al., 2007). In this study the scale had a Cronbach’s alpha 0.97.

#### Health locus of control

We measured health locus of control using the 18-item Form C version of the Multi-dimensional Health Locus of Control (MHLC) measure, developed by Wallston et al. (2005). This is a self-report instrument that assesses the extent to which participants believe their health is attributed to : (1) their own behavior (internality); (2) the behavior of powerful others (which in form C is split into two subscales: doctors and others) or (3) chance, luck, or fate. Participants are asked to rate their agreement with each item using a 6-point Likert-type (1 = strongly disagree; 6 = strongly agree). In this study the scale had a Cronbach’s alpha of 0.75.

#### Resilience scale

Resilience was measured using the Resilience Scale (RS-25) (Wagnild & Young, 1993). It is a self-report with 25 items, each of which is rated on a 7-point Likert scale. Five characteristics of resilience are included in the scale: self-reliance, meaning, equanimity, perseverance, and existential aloneness. RS-25 scores range from 25 to 175. Scores greater than 145 indicate moderately high to high resilience, scores from 116 to 144 indicate low to moderate levels of resilience, and scores of 115 and below indicates very low resilience. In this study the scale had a Cronbach’s alpha of 0.95.

#### Empowerment scale

Empowerment was measured using a 28-item scale questionnaire (Rogers et al., 1997) containing a 4-point agreement scale ranging from 4=strongly disagree to 1=strongly agree. Sample questions from the tool include “I generally accomplish what I set out to do” and “I have a positive attitude about myself”. Total scores ranged from 28 to 112. High levels indicate low levels of empowerment. In this study the scale had a Cronbach’s alpha of 0.88.

#### Ethics considerations

The study was approved by the Research Ethics Committee of Mbarara University and from the Uganda National Council for Science and Technology (# 02/05-20). Adolescents provided assent and/or consent to enrol. For adolescents below the age of consent (below 18 years) we obtained assent after their caregivers provided consent. Emancipated minors (i.e. below 18 years of age but living independently), and empowered adolescents (i.e., below 18 years but responsible for their HIV care per report of their HIV care provider), (Uganda National Council for Science and Technology, 2007) provided their own written informed consent without involvement of their parents/guardians. The RA sought consent from eligible participants after providing and clarifying information. Eligible participants were given a chance to ask questions for clarification prior to giving their consent.

### Data analysis

Descriptive statistics were used to analyze participants’ characteristics and variables associated with internalized HIV stigma including empowerment, resilience, health locus of control and coping self-efficacy. Paired t-test was used to compare median scores of the main predictor variables (health locus of control, coping self-efficacy, resilience, empowerment and internalized HIV stigma) of adolescents attending boarding versus day schools. We also conducted a linear regression model specifying internalized HIV stigma as the outcome variable with empowerment, health locus of control, resilience and coping self-efficacy as the main predictor variables. We also included sociodemographic characteristics (age, sex, boarding school or day school) in the model. All analyses were conducted in Stata version 16 (StataCorp LP, College Station, Texas).

## Results

We recruited 173 participants with average age of 16 (IQR 15-18) years with a minimum and maximum of 13 and 19 years respectively. All participants were single, had formal education and majority had acquired secondary education (66.5%), Table 1.

**Table 1.** Summary characteristics of the participants (n=173).

| **Variables** | **Frequency, n (%)**<br>**or Median (IQR)** |
| --- | --- |
| Mean age (in years) | 16 (15-18) |
| *Sex* |  |
| Male | 76 (43.9) |
| Female | 97 (56.1) |
| *Education level* |  |
| Primary | 47 (27.2) |
| Secondary | 115 (66.5) |
| Tertiary level | 11 (6.3) |
| *School type* |  |
| Boarding | 92 (53.2) |
| Day | 81 (46.8) |
| *Accommodation* |  |
| Rent | 36 (21.4) |
| Own | 135 (78.6) |
| *Food security* |  |
| Yes | 139 (80.4) |
| No | 34 (19.6) |
| *Current caregiver* |  |
| Both parents | 54 (31.2) |
| Mother alone | 71 (41.0) |
| Father alone | 7 (4.1) |
| Siblings | 9 (5.2) |
| Grandparents | 23 (13.3) |
| Other relatives | 9 (5.2) |
| HIV stigma score | 3 (2-4) |
| Health locus of control score |  |
| Internal | 30 (24-35) |
| Chance | 23 (19-26) |
| Powerful others | 31 (28-33) |
| Resilience | 131 (110-145) |
| Coping self-efficacy | 155 (125-193) |
| Mean empowerment | 63 (59-67) |

\* Other relatives; aunt=5, guardian =1, uncle=1, stepmother=1 alone = 1
S.D = Standard Deviation.

### Intrapersonal factors

The internalized HIV stigma scores were significantly higher among adolescents attending boarding schools compared to their peers in day schools. There were also statistically significant differences in the mean scores of internalized HIV stigma and intrapersonal factors (health locus of control, resilience, empowerment and self-efficacy) between participants who were in boarding schools and those who were in day schools (Table 2).

**Table 2.** Comparing mean scores of Intrapersonal factors of the participants stratified by type of school.

| Variable | School type |  |  |
| --- | --- | --- | --- |
|  | Mean (SD) |  | p value |
|  | Boarding | Day |  |
| Internalized AIDS-related stigma | 3 (1.5) | 2.6 (1.4) | **<0.001** |
| Health locus of control score |  |  |  |
| Internal | 27.3 (7.4) | 30.4 (6.2) | **0.002** |
| Chance | 22.8 (5.2) | 22.2 (5.8) | 0.236 |
| Powerful others | 31.4 (3.6) | 29.3 (4.1) | **<0.001** |
| Resilience score | 123.2 (22.9) | 135.0 (20.2) | **<0.001** |
| oping self-efficacy score | 146.0 (40.6) | 171.4 (38.3) | **<0.001** |
| Mean empowerment score | 63.4 (6.4) | 61.7 (6.2) | **0.036** |

s.d = standard deviation

### Intrapersonal factors associated with Internalized HIV stigma

After adjusting for covariates the linear regression model showed that internal health locus of control (b=-0.08, C.I: -0.11- -0.05), resilience (b=-0.03, C.I: -0.4- -0.02) and coping self-efficacy (b=-0.02, C.I: -0.02- -0.01) were negatively correlated with internalized HIV stigma while empowerment (b=0.05, C.I: 0.02-0.090) was positively correlated. However, after mutually adjusting for the main predictors (intrapersonal factors) and socio-demographic factors, only internal locus of control (b=-0.05, C.I: -0.08- -0.01) and coping self-efficacy (b=-0.02, C.I: -0.02- -0.01) remained statistically significantly correlated with HIV stigma (Table 3).

**Table 3.** Association between internalized stigma HIV stigma and intrapersonal factors (internal locus of control, resilience, coping self-efficacy and empowerment).

| Variable | Separately adjusted for covariates | Mutually Adjusted each other |
| --- | --- | --- |

|  | b (95% CI) | P-value | b (95% CI) | P-value |
| --- | --- | --- | --- | --- |
| Health locus of control |  |  |  |  |
| Chance | 0.01 (-0.03-0.05) | 0.50 | 0.03 (-0.01-0.06) | 0.11 |
| Internal | -0.08 (-0.11- -0.05) | **<0.001** | -0.05 (-0.08- -0.01) | **0.008** |
| Powerful others | 0.01 (-0.04- -0.07) | 0.65 | 0.01 (-0.04-0.06) | 0.59 |
| Resilience | -0.03 (-0.4- -0.02) | **<0.001** | 0.01 (-0.01-0.02) | 0.44 |
| Empowerment | 0.05 (0.02-0.090) | **0.003** | 0.02 (-0.02-0.05) | 0.29 |
| Coping self-efficacy | -0.02 (-0.02- -0.01) | **<0.001** | -0.02 (-0.02- -0.01) | **<0.001** |

## Discussion

The study findings showed higher levels of internalized HIV stigma among ALHIV in boarding schools compared ALHIV in day schools. Additionally, ALHIV in boarding schools were less resilient and with lower scores on the coping self-efficacy scale. However, ALHIV in boarding schools scored higher on the empowerment scale. The study also found that internal health locus of control, resilience and coping self-efficacy had a negative correlation with internalized HIV stigma while empowerment scores had a positive correlation. However after mutually adjusting for the predictors, internal locus of control and self-efficacy retained negative correlation with stigma.

These findings are consistent with those of a study conducted in southwestern Uganda, which found that ALHIV students attending boarding schools reported feeling stigmatized in addition to a higher risk of bullying following unintentional HIV status disclosure. (Kihumuro et al., 2021). HIV stigma affects the adolescents’ability to adhere to their ART medication as they struggle to keep their HIV status a secret (Ashaba et al., 2018; Inzaule et al., 2016; Kihumuro et al., 2021; Mutwa et al., 2013; Zanoni et al., 2019). Unintended disclosure of their HIV status may aggravate HIV stigma and bullying (both physical and verbal abuse), resulting in a lack of social support that negatively impacts ART adherence, while others drop out of school to remain in HIV care. (Ashaba et al., 2019; Jacobi et al., 2020; Kihumuro et al., 2021; Kimera et al., 2020). This is further worsened by the developmental stage where adolescents are beginning to form identity and value their peer relationships as they grow toward maturity and independence since these are important immediate sources of social and emotional support in boarding school (Folayan et al., 2014). The school environment does not offer the necessary privacy to enable the ALHIV take their HIV medicines (Abubakar et al., 2016; Wolf et al., 2014). Moreover adolescents in boarding schools lack the support from parents while they are away at school yet the school authorities do not offer the necessary support (Mutwa et al., 2013). Furthermore, school environments are unfavorable due to strict routines that require ALHIVs to ask for permission to honor their clinical appointments for medical refills and reviews, which increases the risk of HIV status exposure and thus HIV stigma, while the time spent traveling from school to the clinic causes the majority of students to fall behind in their academics and increases the risk of bullying (MacCarthy et al., 2018; Zanoni et al., 2019).

The finding that internal health locus of control, resilience, and coping self-efficacy had a negative correlation with internalized HIV stigma is in line with studies which have documented that poor coping self-efficacy and low internal health locus of control are associated with high internalized stigma among both adolescents and adults which results into feeling less empowered (Andren et al., 2011; Andrinopoulos et al., 2010; Kurniawan & Fitrio, 2019). Conversely, high internal locus of control is associated with positivity, and the feeling of being in charge of one’s health hence less feelings of stigmatization and good health outcomes such as adherence to medications (Basinska & Andruszkiewicz, 2012; Nazareth, 2016). Also adolescents who feel well empowered are less likely to feel stigmatized because they have the right knowledge and understand their condition well hence can make well-informed decisions knowing the likely consequences as opposed to those who are less empowered (Náfrádi et al., 2017). This highlights the need to empower ALHIV with the right knowledge about their condition, treatment, possible side effects of the medication as well as disclosure, so that they can make well informed health related decisions and hence reducing internalized stigma. This should involve developing adolescent friendly home/community as well as school based programs aimed at empowering ALHIV as recommended by the World Health Organization (Soeters et al., 2020; World Health Organization, 2015). Unfortunately, there is a disparity in access to this information between day and boarding students, necessitating the need to ensure that the designed programs can be effectively implemented in both contexts. By empowering and supporting these adolescents, they will gain control of their condition and also have higher coping self-efficacy, which will improve their resilience and treatment outcomes (Earnshaw et al., 2015). As a result, higher resilience results in reduced emotions of stigmatization among adolescents, which improves treatment outcomes such as ART adherence (Gottert et al., 2019; Harrison et al., 2019). Therefore, the findings of this study highlight the need to design empowering interventions aimed at promoting self-efficacy, internal health locus of control and resilience among ALHIV in order to reduce internalized stigma. This will have far reaching effects in improving adherence to treatment resulting in better treatment outcomes and quality of life among adolscents living with HIV.

However, this study was conducted in a health facility where all children involved were on ART treatment hence may not clearly represent community setting where some children may not be on treatment. There is also need to assess the effect of these factors among children with vertically versus horizontally acquired HIV since this has significant psychosocial implication.

## Conclusion

The study findings show that ALHIV attending boarding schools have higher levels of HIV stigma compared to their peers in day schools and that stigma is associated with low resilience, coping self-efficacy, empowerment and internal health locus of control. Therefore, boarding schools need more sensitization regarding HIV/AIDS to reduce on the bullying, stigmatization and dicrimination of adolescents with HIV but instead provide a more supportive environment that will improve their resilience, coping self-efficacy and empowerment. The findimgs highlight the role of intrapersonal factors in fluencing stigma among adolescents livng with HIV. Hence there is need to design interventions focusing on internal locus of control, self efficacy, empowerment and resilience among adolescents in order to reduce internalised HIV stigma which in turn will improve treatment outcomes.

## Data Availability

All relevant data are within the manuscript and its Supporting Information files.

## Acknowledgement

Special appreciation goes to participants who voluntarily gave useful information that has yielded this manuscript which will contribute to science as well as the people who collected this data.

